# Human genetic factors associated with pneumonia susceptibility, a cue for COVID-19 mortality

**DOI:** 10.1101/2021.06.03.21258106

**Authors:** Debleena Guin, Saroj Yadav, Priyanka Singh, Pooja Singh, Sarita Thakran, Samiksha Kukal, Neha Kanojia, Priyanka Rani Paul, Bijay Pattnaik, Viren Sardana, Sandeep Grover, Yasha Hasija, Indian Genome Variation Consortium, Anurag Agrawal, Ritushree Kukreti

## Abstract

The risk for community acquired pneumonia (CAP) is partially driven by genetics. To identify the CAP-associated genetic risk loci, we performed a meta-analysis of clinically diagnosed CAP (3,310 individuals) with 2,655 healthy controls. The findings revealed *CYP1A1* variants (rs2606345, rs4646903, rs1048943) associated with pneumonia. We observed rs2606345 [G vs T; OR=1.49(1.29-1.69); p=0.0001; I^2^= 15.5%], and rs1048943 [T vs G; OR= 1.31(0.90-1.71); p=0.002; I^2^=19.3%] as risk markers and rs4646903 [T vs C; OR= 0.79(0.62-0.96); p=0.03; I^2^=0%] as a protective marker for susceptibility to CAP, when compared with healthy controls. Our meta-analysis showed the presence of *CYP1A1* SNP alleles contributing significant risk toward pneumonia susceptibility. Interestingly, we observed a striking difference of allele frequency for rs2606345 (*CYP1A1*) among Europeans, Africans and Asians which may provide a possible link for observed variations in death due to coronavirus disease 2019 (COVID-19), a viral pneumonia. We report, for the first time, a significant positive correlation for the risk allele (T or A) of rs2606345, with a higher COVID-19 mortality rate worldwide and within a genetically heterogeneous nation like India. Mechanistically, the risk allele ‘A’ (rs2606345) is associated with lower expression of *CYP1A1* and presumably leads to reduced capacity for xenobiotic detoxification. We note that ambient air pollution, a powerful inducer of *CYP1A1* gene expression, is globally associated with lower, not higher mortality, as would normally be predicted. In conclusion, we find that *CYP1A1* alleles are associated with CAP mortality, presumably via altered xenobiotic metabolism. We speculate that gene-environment interactions governing *CYP1A1* expression may influence COVID-19 mortality.

## Introduction

The ongoing coronavirus disease 2019 (COVID-19) pandemic and consequent mortality have rarely been examined through the lens of community acquired pneumonia (CAP). Pneumonia is an inflammatory condition of the lungs usually caused by bacterial or viral infection. According to the site of acquisition, pneumonia is classified as community acquired (CAP) or nosocomial (NP). The global burden of disease study (2015) stated that lower respiratory infections like pneumonia are the third most common cause of death globally [1]. Pneumonia occurs when pathogens enter in the alveoli, infect, multiply and encourage a host immune response. These responses cause inflammation of the lung tissues, marking the pathogenesis of pneumonia [2]. Host genetic factors that participate in these processes starting from pathogen entry, infection, inflammation, and resolution can all be considered as good candidates in genetic association studies of pneumonia and its complications [3, 4].

A number of studies have investigated the genetic association as well as gene interaction of pneumonia risk with monooxygenase enzyme group, cytochrome P450 (CYP). In a study, a group of researcher identified CYP1A1 cytochrome P450 family 1 subfamily A member 1 (*CYP1A1*) as a critical regulator of inflammatory responses and phagocytosis in sepsis some through *CYP1A1*-involved signalling pathways that may be promising targets for treating sepsis or other inflammatory diseases [5]. Interestingly, few other studies indicated towards the role of *CYP1A1* genetic polymorphisms in infectious diseases and consequently establishing its role in inflammatory responses. Previously, it was identified that genetic variants of some host genes (*CYP1A1, ACE* and *IL-6*) are associated with the diversity in response to CAP [6, 7]. The selection of this gene was established based on its role in physiological and pathological processes during pneumonia infection, particularly in the immune and inflammatory responses [6]. Hence, we selected this gene. The most widely reported *CYP1A1* single nucleotide polymorphisms (SNPs) were rs2606345, rs4646903 and rs1048943. These SNPs had functional consequences which may ultimately be involved with a disease phenotype like pneumonia. The intron-located rs2606345 (C>A) determines enhanced gene expression in the presence of specific substrates (allele C) or in their absence (allele A). The minor allele of rs4646903C, located near the 3’ UTR, also shows an increased inducibility. Another *CYP1A1* SNP rs1048943 (T>A, C, G) (Ile/Val), resulted in a missense amino acid substitution, is characterized by the substrate-specific increased activity for minor allele G. Thus we can suggest that genetically determined alteration of *CYP1A1* activity could contribute to lung inflammation pathogenesis. Thus, while there may be several evidence directing towards the role of this gene and/or its genetic variants in pneumonia susceptibility, so far, we lack a common mechanistic framework that integrates it all.

Here, we report a meta-analysis of pneumonia from ten included studies. We investigated the association of the *CYP1A1* gene and its genetic variants (rs2606345, rs4646903, rs1048943) with the risk of pneumonia, both CAP and NP, independently for all samples. The primary objectives of this meta-analysis are: (1) to determine a precise numerical estimate of the impact of *CYP1A1* risk allele and pneumonia risk, (2) to determine whether age or sex alters the odds of incident pneumonia in presence of the *CYP1A1* SNP alleles (3) to determine whether any association between *CYP1A1* and CAP is generalizable to COVID-19. We hypothesized that *CYP1A1* genetic variant would increase the odds of incident pneumonia and COVID-19 mortality.

## Results

The current systematic search included eleven studies totalling 5,965 unique subjects (3,310 cases and 2655 healthy controls). The flow chart for the study selection process is represented in **Figure 1**. Demographic characteristics and clinical details of all the included studies are provided in **Supplementary table 1**. The study populations primarily comprised of well-characterized cohorts in Russia [7-15] and one included Chinese cohort [6]. The mean age of all the pooled participants was 29.33 years (31.16 years for cases and 27.49 years for controls). All the studies discuss the association of *CYP1A1* genetic variants (rs2606345, rs4646903, rs1048943) with the risk of pneumonia. For cumulative quality assessment, three of ten articles were deemed as good quality (cut off score of ≥7), six articles (≥5–6 score) were categorised under moderate, and finally any scores below 5 were judged as poor quality which included one article **(Supplementary table 2)**.

**Figure 1:**
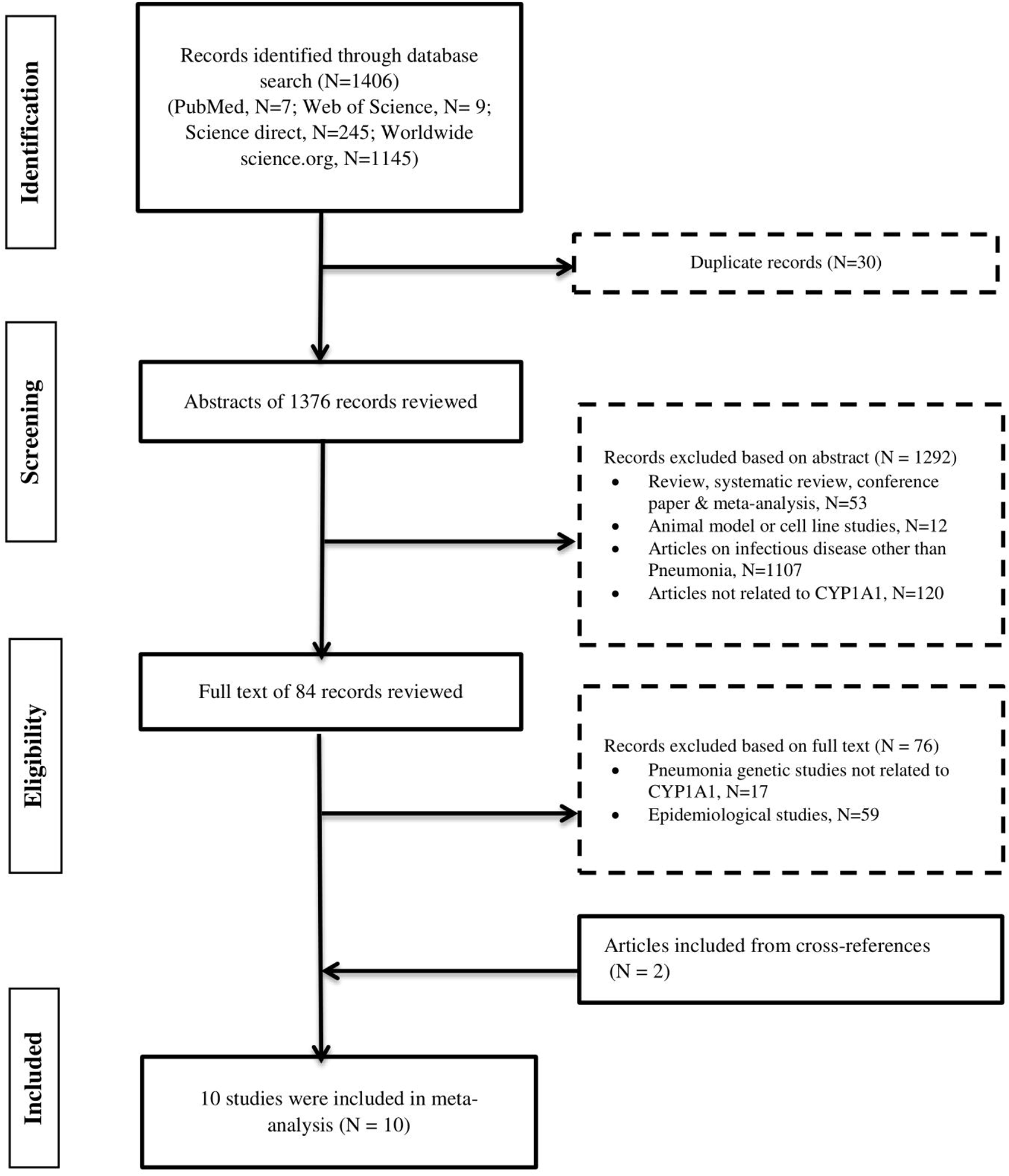
Flow chart of study selection in metaanalysis of CYP1A1 polymorphisms with Pneumonia risk.Study methodology for the inclusion and exclusion of studies exploring the role of *CYP1A1* genetic variants in pneumonia patients. The number of studies excluded on each step is represented as N.

This meta-analysis has been categorised into two types of pneumonia, CAP and NP, based on the cases included in each study (**Table 1**). Among all the ten studies included comparing patients with pneumonia with healthy controls, ten provided the data on rs2606345, ten provided data on rs1048943 and eight provided the data on rs4646903. Of all the *CYP1A1* variants studied, we observed the most significant distribution of rs2606345, rs4646903, and rs1048943, with data available for 1,336, 1,382, and 909 individuals with CAP and 632, 508, and 449 individuals with NP, respectively. Overall, statistically significant heterogeneity (P<0.10, I^2^<50%) was observed among these included studies, therefore the fixed effect models were used for the following analysis. Our meta-analysis demonstrated that *CYP1A1* genetic polymorphisms significantly correlated with the increased risk of pneumonia under the allele model rs2606345 [G vs T; OR=1.49(1.29-1.69); p=0.0001; I^2^= 15.5%], and rs1048943 [T vs G; OR= 1.31(0.90-1.71); p=0.002; I^2^=19.3%] as risk markers and rs4646903 [T vs C; OR= 0.79(0.62-0.96); p=0.03; I^2^=0%] as a protective marker for susceptibility to CAP, when compared with healthy controls.

**Table 1:** Pooled odds ratio for allelic comparisons for studies exploring association of *CYP1A1* variants-rs2606345, rs4646903, rs1048943 in patients with risk of CAP.

| Gene (SNP) | Risk allele | No of studies | Population | Total samples | CAP patients |  | Total | Control |  | Total | OR(95%CI) | P value | I<sup>2</sup> | Model | Test of publication bias |
| --- | --- | --- | --- | --- | --- | --- | --- | --- | --- | --- | --- | --- | --- | --- | --- |
|  |  |  |  |  | Risk allele present | Risk allele absent |  | Risk allele present | Risk allele absent |  |  |  |  |  | Egger's test |
| *CYP1A1* (rs2606345) | T | 5 | Chinese, Russian | 2691 | 2112 | 560 | 2672 | 1876 | 834 | 2710 | 1.49(1.29-1.69) | **0.0001** | 18.88 | F | 0.361 |
| *CYP1A1* (rs4646903) | C | 3 | Russian | 1929 | 168 | 1650 | 1818 | 231 | 1809 | 2040 | 0.79(0.62-0.96) | **0.03** | 0.00 | F | 0.948 |
| *CYP1A1* (rs1048943) | G | 6 | Russian | 2469 | 111 | 2007 | 2118 | 99 | 2721 | 2820 | 1.31(0.90-1.71) | **0.002** | 0.00 | F | 0.002 |

**Bold** characters highlight significantly associated alleles with respective P values. CAP, community acquired pneumonia; OR, odds ratio; CI, confidence interval; F, Fixed effect model. All p values calculated using chi-square test

### Community acquired pneumonia (CAP)

The pooled OR derived from 3,310 cases and 2,655 healthy controls subjects in 10 recruited studies was statistically significant (see **Table 1**). Homogeneity analysis for the ORs from the ten studies of the risk allele suggested that there was statistically significant evidence for heterogeneity of the odds ratio (ORs) among the other groups. The genetic variant, rs2606345 and rs1048943 were significantly overrepresented in CAP patients with ORs of 1.49(1.29-1.69); p=0.0001, and 1.31(0.90-1.71); p=0.002; respectively and qualified as risk markers (**Figure 2 A** and **C**). In contrast, rs4646903 was observed to be protective (0.79(0.62-0.96); p=0.03) (**Figure 2 B**).

**Figure 2:**
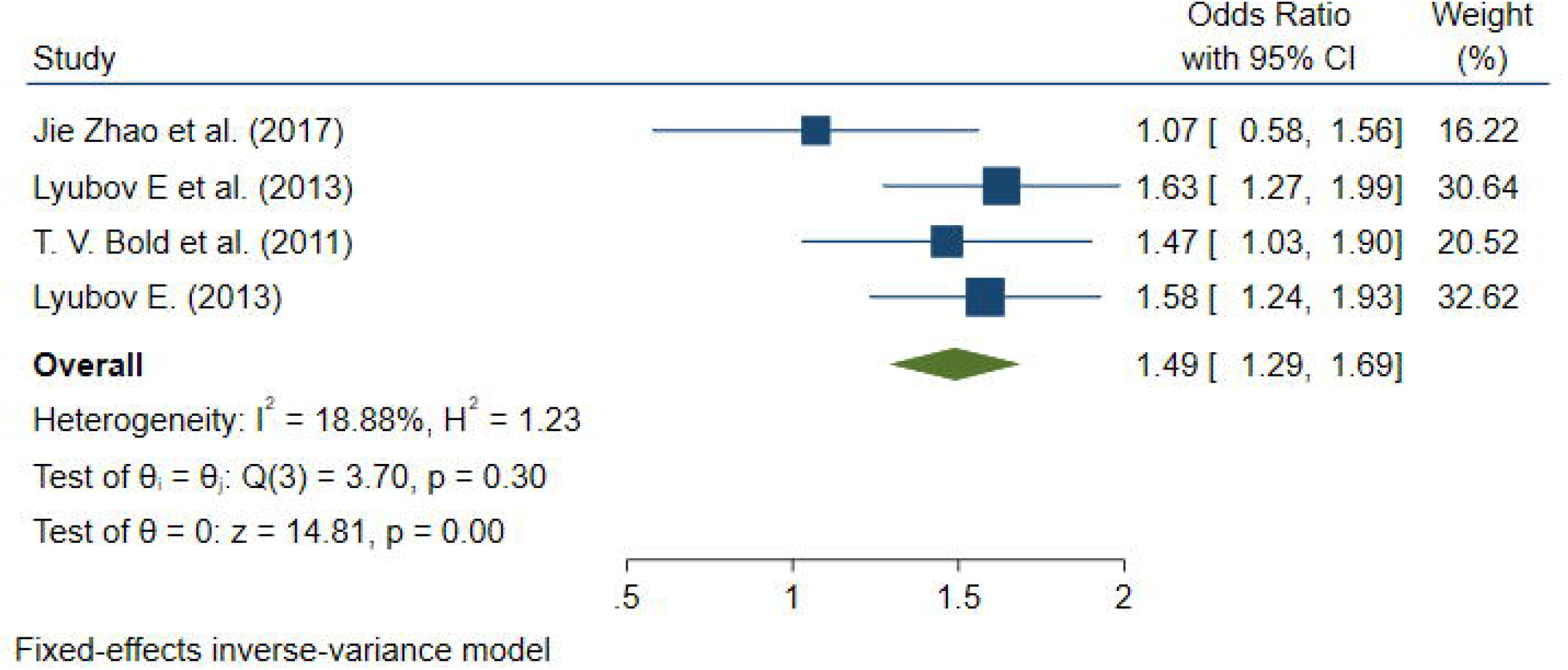
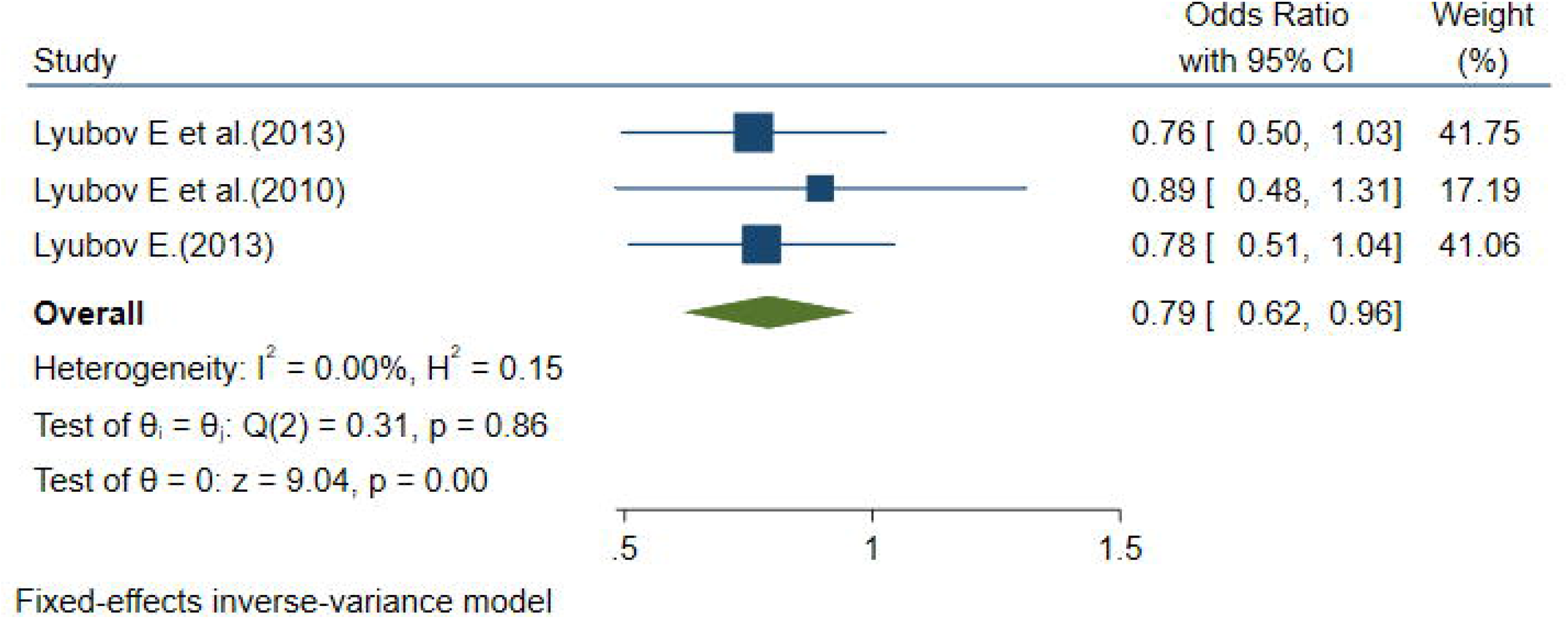
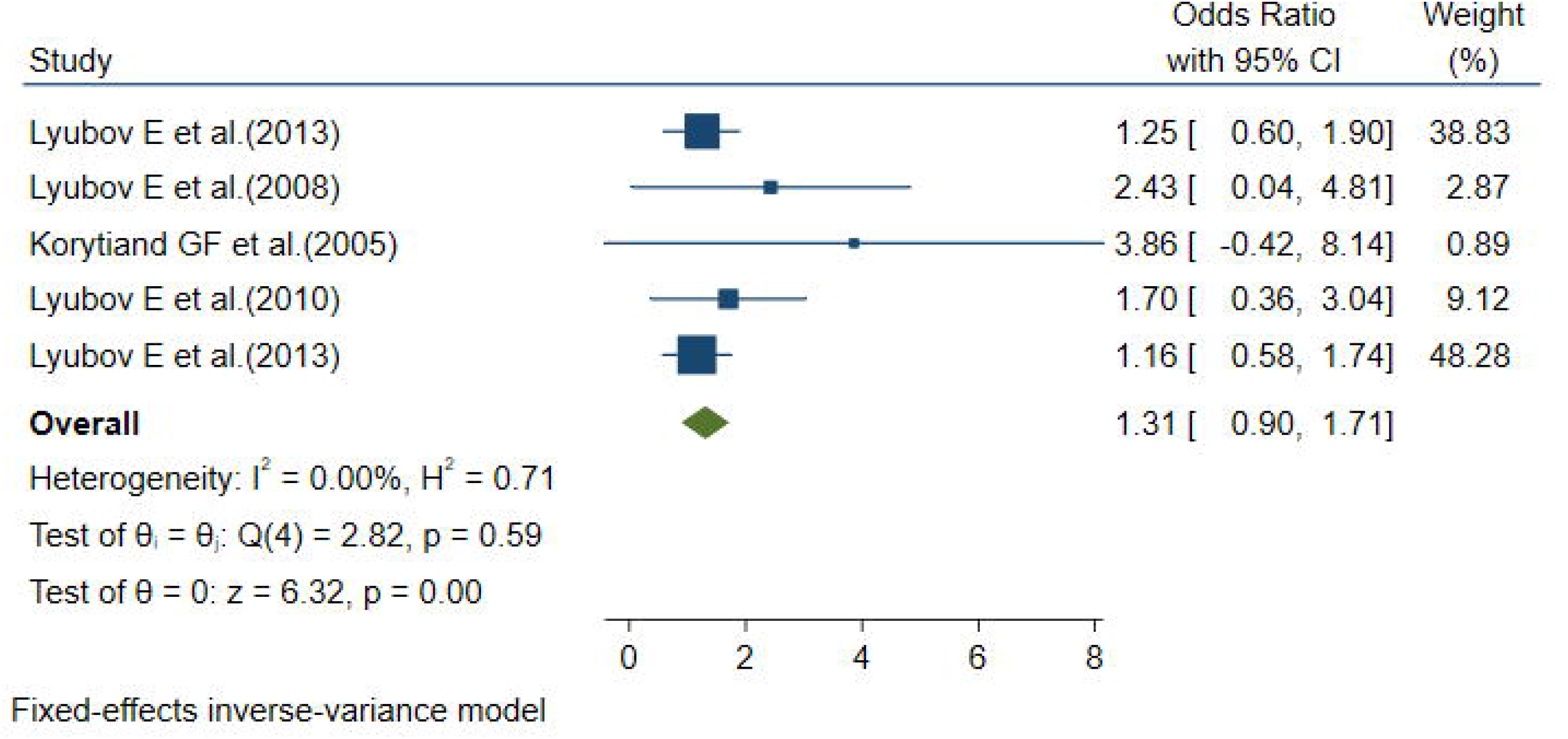
Forest plot determining pneumonia risk associated of *CYP1A1* genetic variants: a) for rs2606345, b) rs4646903 c)1048943; in community acquired pneumonia (CAP). The square and horizontal lines correspond to the study-specific odds ratio(OR) and 95% confidence interval (CI). The area of the square refers to the study specific weight (inverse of variance).The diamond represents the summary of OR and 95% CI.

### Nosocomial pneumonia (NP)

The pooled OR derived from 3,310 cases and 2,655 healthy controls subjects in ten recruited studies was not statistically significant (**Supplementary table 1**). The risk allele of all the three *CYP1A1* genetic variants were seen to have increased frequency in NP patients as compared to healthy controls, however no statistically significant correlation was identified between the polymorphisms and the risk of NP..

### Test for publication bias

We failed to observe publication bias in any of the associations reported above, showing that our results were reliable (**Table 1 and Supplementary figure 1**).

### Genetic variability of rs2606345 in global populations and a possible link to COVID-19

Since rs2606345 is the major allele in Europeans (66.6%) but not in other populations (African 5%, Asian 5-30%) [16], we explored a possible association with high variability in regional mortality of the ongoing COVID-19 pandemic, through a spatial analysis of the association between COVID-19 mortality with the alternate allele of rs2606345 (*CYP1A1*). To estimate the correlation pattern we performed a linear model regression and Pearson’s correlation coefficient exploring the variation of COVID-19 mortality with the allele frequency data (for rs2606345) for each country (**Figure 3, A** and **B**). The regression analysis showed a strong positive correlation (p <0.22 × 10^−16^) of higher mortality with higher frequency of A (or T), with a large effect size goodness of fit (r^2^) ∼ 43.5% (**Figure 3 D**). The spatial analysis in the genetically heterogeneous Indian population also confirmed a similar trend (**Figure 4 A** and **B**), with regions that have European genetic ancestry being more affected [17, 18]. The linear model analysis revealed a positive correlation between mortality related to COVID-19 and alternate allele frequency (p <0.039, r^2^ =18.8%) (**Figure 4 C**).

**Figure 3:**
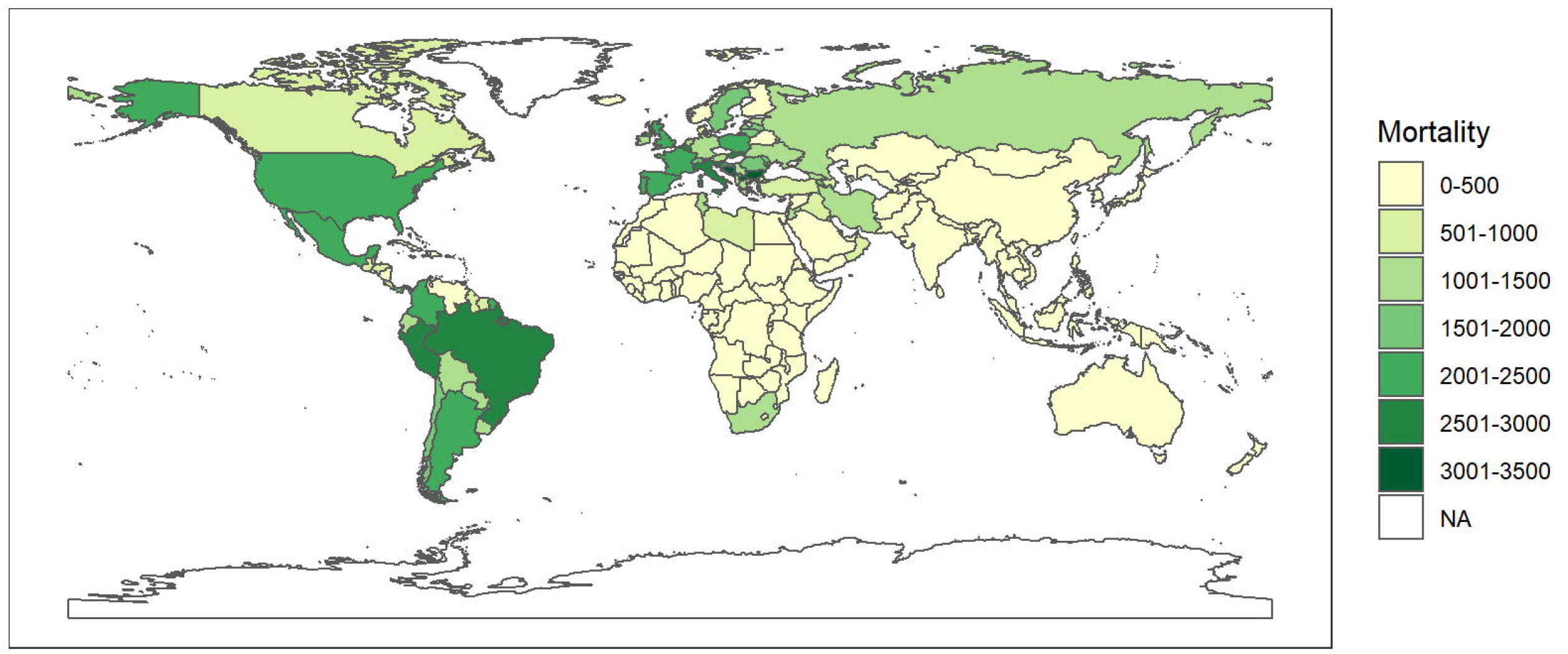
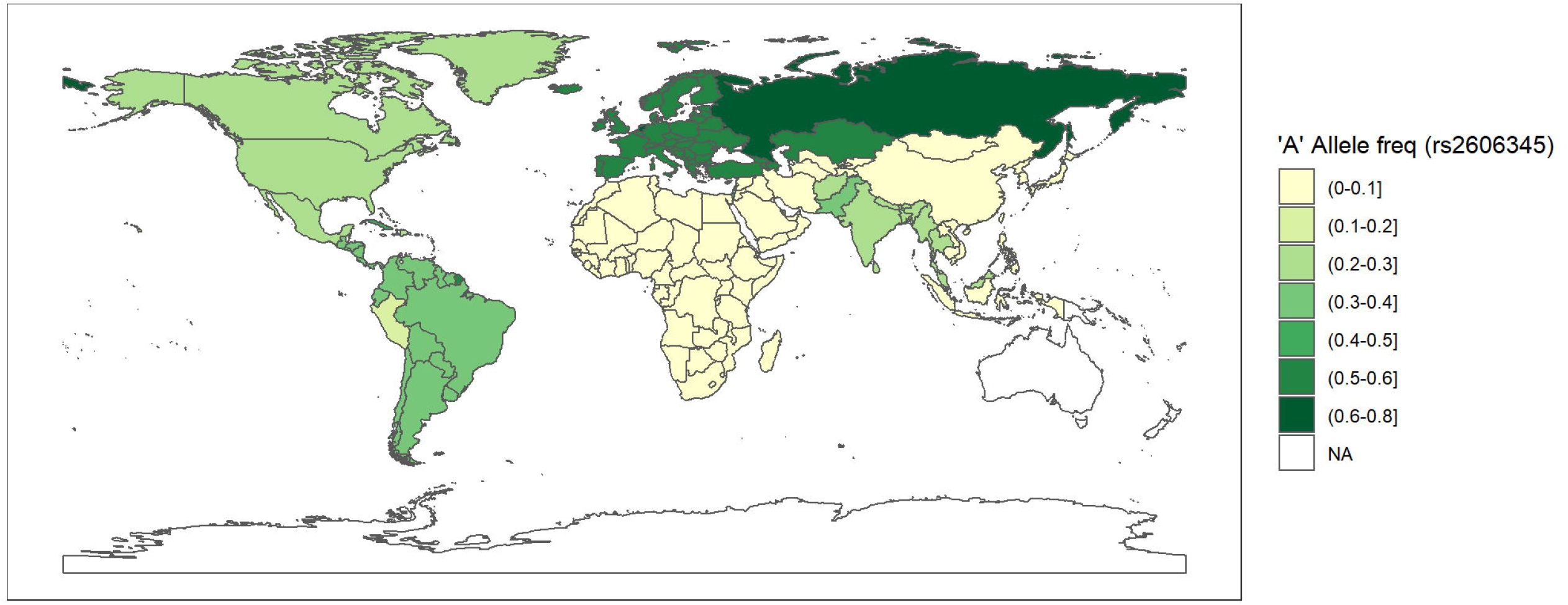
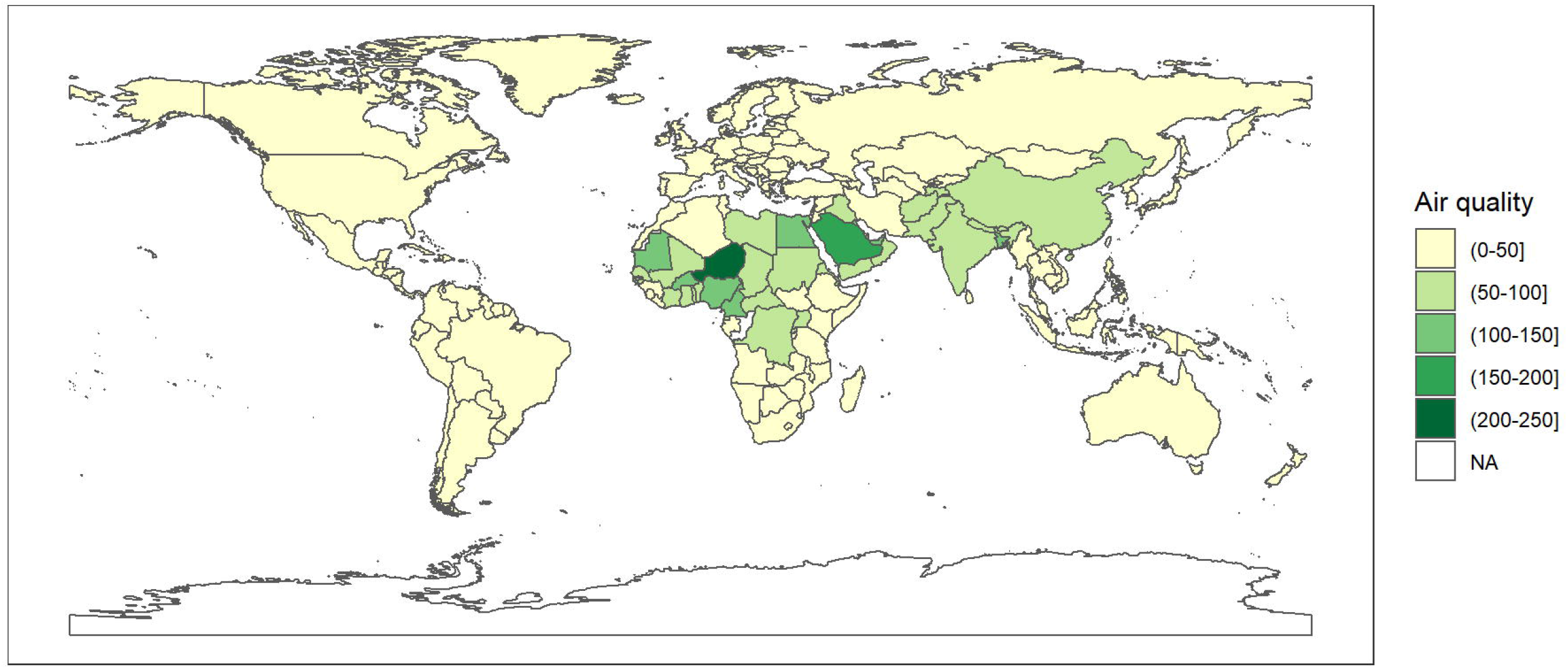
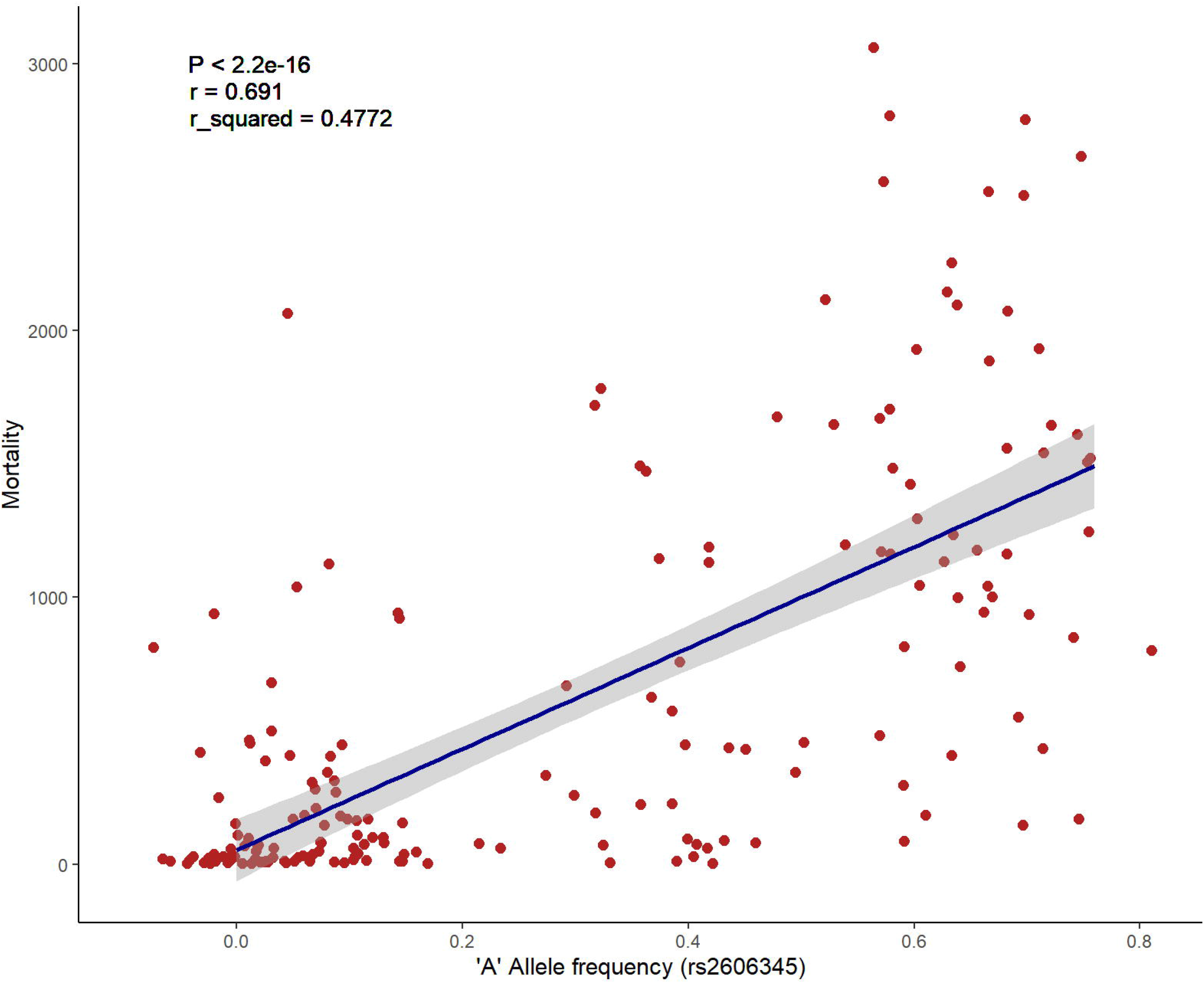
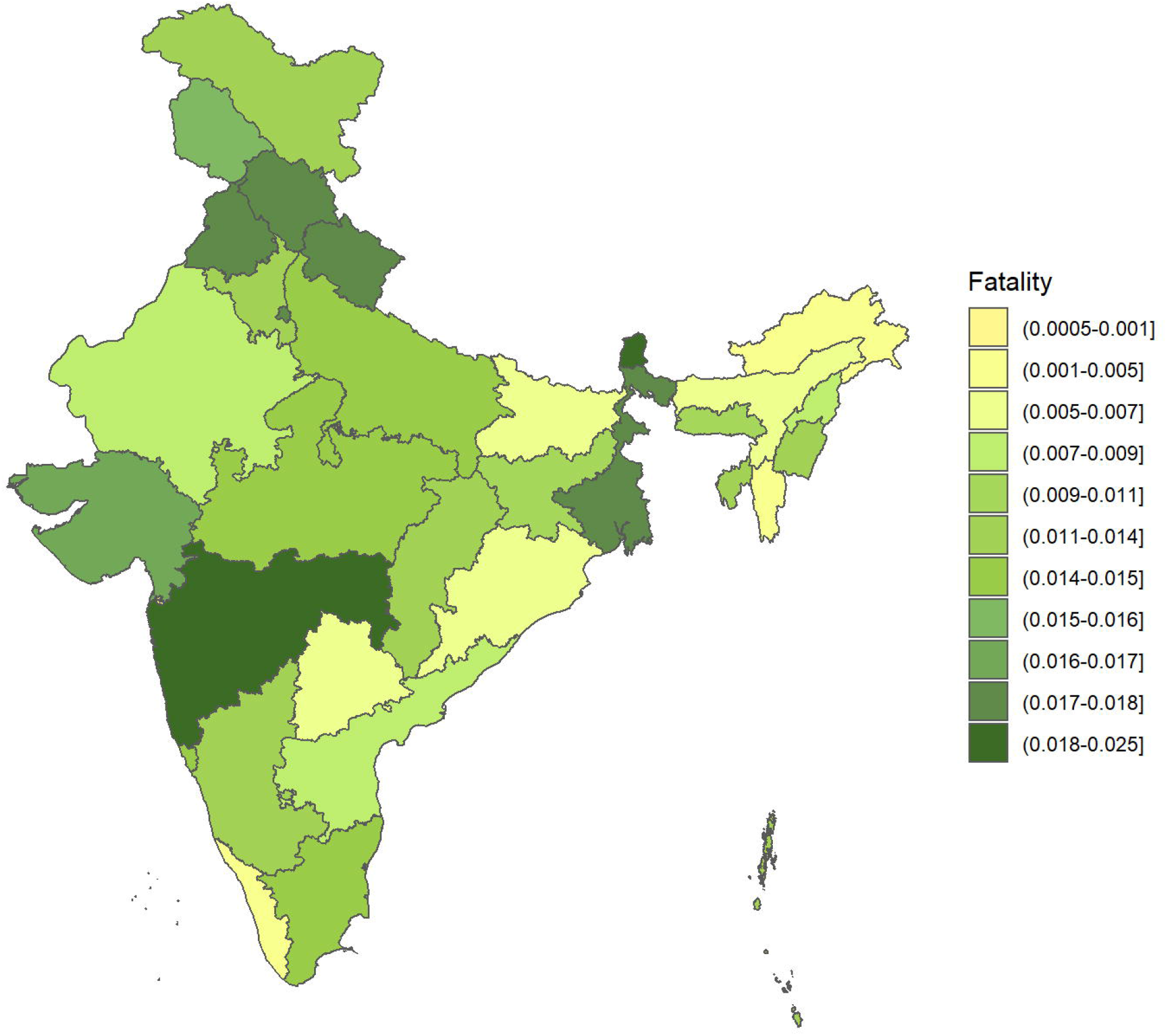
A contrasting feature between allele frequency of rs2606345 (*CYP1A1*), COVID-19 mortality rate, and particulate matter air pollution worldwide. **A**. Geospatial frequency maps depicting the distribution of minor allele frequency of rs2606345 (*CYP1A1*) SNPs. **B**. A geospatial density map of the number of deaths per million inhabitants as on 24 May 2021 worldwide due to COVID-19. **C**. A geospatial density map of the number of air quality index (AQI) PM_2.5_ worldwide obtained from ourworldindata.org till 2016. **D**. The regression analyses showing the goodness of fit and Pearson correlation coefficient for the allele frequency and COVID-19 death per million. Some of the values have been jittered by drawing samples from uniform distribution. **E**. The regression analyses showing the goodness of fit and Pearson correlation coefficient for the AQI PM_2.5_ and COVID-19 case fatality ratio across Indian states. COVID-19 mortality is extracted from ourworldindata.org till 24 May 2021. The minor allele frequency distribution is plotted for SNPs across global populations of rs2606345. The data is obtained from population frequency data of the 1000genome browser on 8 March, 2021. Air quality is represented by concentration of particulate matter of size 2.5 micrometres (µm) (PM_2.5_). The data is obtained from ourworldindata.org till 2016. The white coloured areas in the map show the absence of data. A half open intervals includes only one of its end-points and is denoted by mixing notations for open and closed intervals. For e.g., (0-1] means greater than 0 and less than or equal to 1 and [0,1) means greater than or equal to 0 and less than 1.

**Figure 4:**
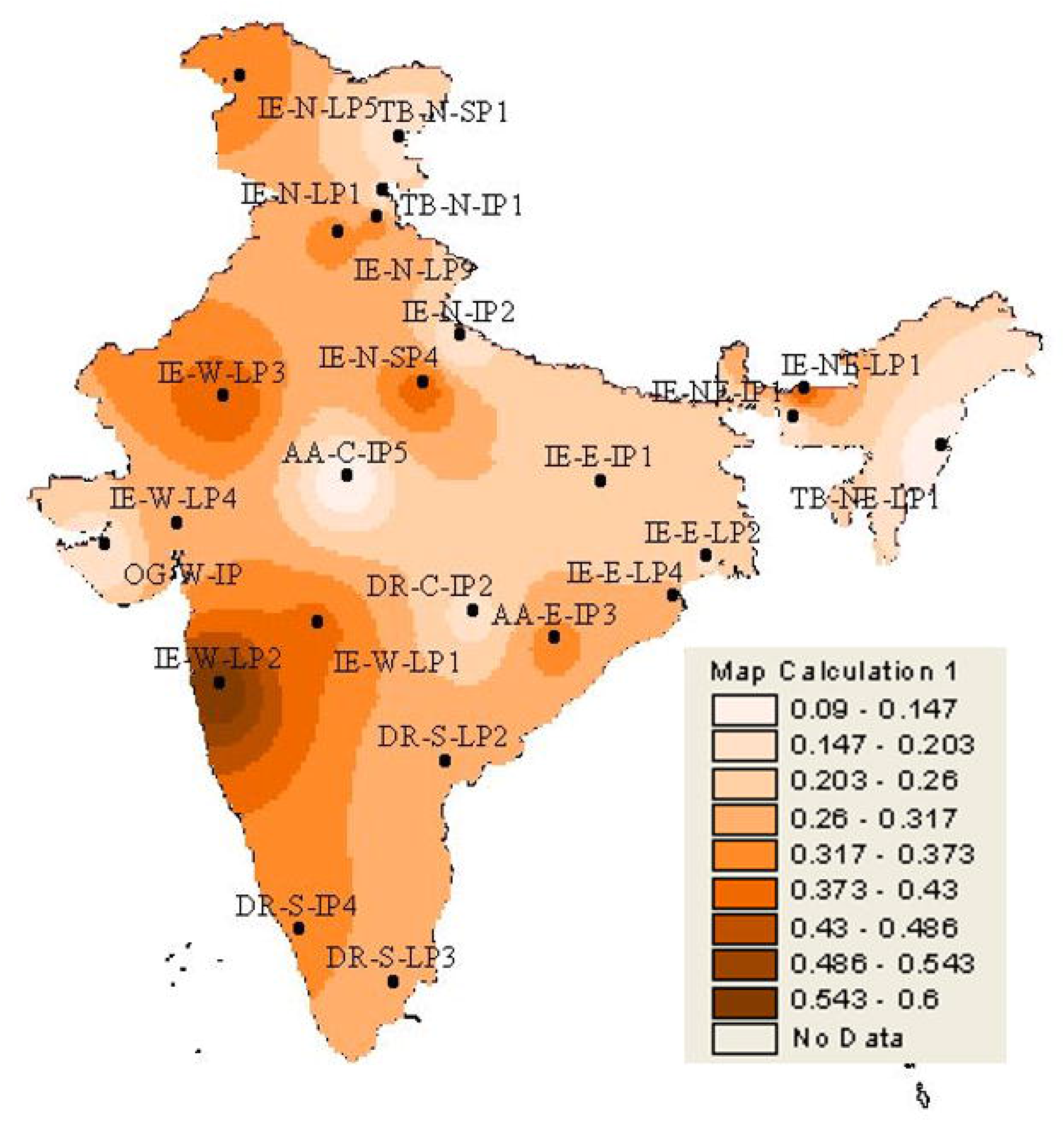
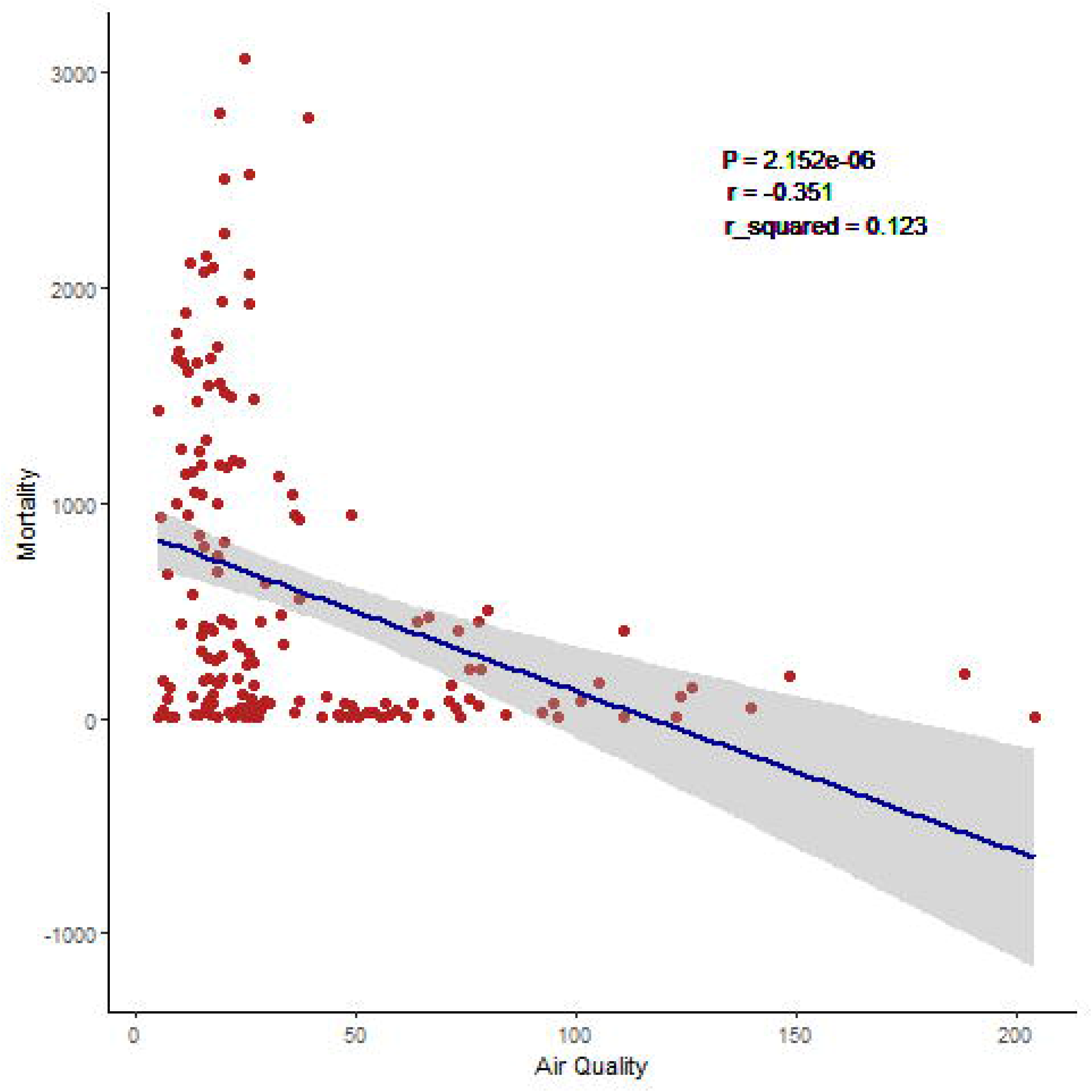
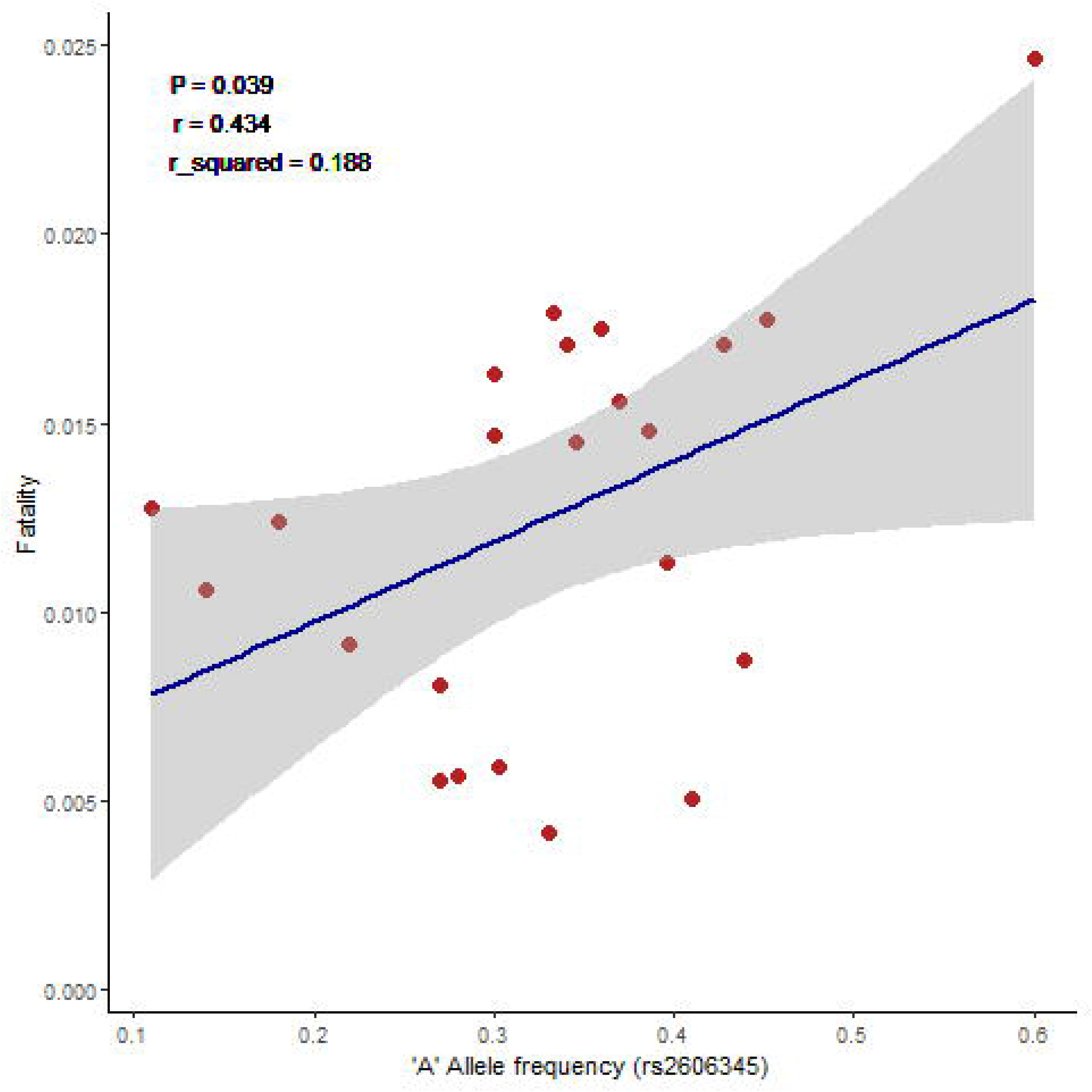
A contrasting feature between COVID-19 mortality rate and allele frequency data of rs2606345 (*CYP1A1*) in Indian population. **A**. Indian population state-wise frequency of case fatality rate (CFR) for COVID-19. CFR data given till 24 May, 2021. **B**. Allelic ‘A’ distribution of rs2606345 variant in major ethnic groups in the Indian population. **C**. The regression analyses showing the goodness of fit and Pearson correlation coefficient for the allele frequency and COVID-19 case fatality ratio across Indian states. The COVID-19 case fatality ratio has been extracted from MoHFW.gov.in. Here, CFR is calculated as number of death upon number of death + number of recovered from COVID-19 infection, since COVID-19 is an ongoing pandemic [60]. A half open intervals includes only one of its end-points and is denoted by mixing notations for open and closed intervals. For e.g., (0-1] means greater than 0 and less than or equal to 1 and [0,1) means greater than or equal to 0 and less than 1. Figure 4b reproduced with permission [17].

Additionally, air pollution is known to upregulate *CYP1A1* expression [19], similar to what the protective allele is expected to do, we estimated the correlation between the population-weighed average level of exposure to concentration of suspended particulate matter in air measuring less than 2.5 microns in diameter (PM_2.5_) (**Figure 3 C**) with COVID-19 mortality. We observed an inverse relationship (r=-0.35) between the two. **Figure 3 E** represents the significant negative correlation (p < 2.15 × 10^−6^) with the goodness of fit (r^2^) estimated to be 12.3%. This suggests additional gene-environment interactions. In view of the preliminary nature of the evidence, this was not explored further at this point.

## Discussion

This study is an attempt to investigate the role of host genetic factors in pneumonia susceptibility. Like pneumonia, most infectious diseases, are a consequence of a complex network of host genetics and pathogen genetic factors that may be inducible by several non-genetic factors. The host immune mediated response determines the course of the disease, its susceptibility, progression, and severity. Such immune response related genes may serve as good candidates in establishing a genetic association in infectious diseases. Therefore, we considered xenobiotic detoxification genes encoding, CYP enzymes, which are also involved in inflammatory responses by inducing oxidative stress on encountering foreign bodies[20]. This meta-analysis reiterates prior observations that suggest a genetic contribution of *CYP1A1* to pneumonia risk, and provides a more precise estimate of the risk of individuals carrying *CYP1A1* genetic variants (rs2606345, rs4646903, rs1048943) for developing pneumonia.

Our study revealed that the alternate allele (A in plus-strand or T in minus-strand) of rs2606345 increased pneumonia susceptibility in Russian population. We also observed, this allele to be the major allele in European (66.6%) and Russian (∼80%) population unlike in the other populations (African 5%, Asian 5-30%, American 39%) [16]. Interestingly, we observed a striking similarity in the trend for the recent outbreak of COVID-19, a global pandemic. We would also like to warn that since this is an ongoing pandemic, the numbers are changing with time and this is circumstantial evidence. With the second surge progressing, we still observe similar trends with higher COVID-19 mortality in populations with higher A allele frequency (rs2606345) (**Figure 3 B** and **D**) Likewise, the spatial analysis in the Indian population also directed towards a similar trend, where the frequency of alternate allele of this SNP (rs2606345) varied from 9% (in sub-population of northern Madhya Pradesh) to 60% (in sub-population of Maharashtra) [17] (**Figure 4 A** and **B**). The highest number of COVID-19 deaths have been observed in the west-Indian state of Maharashtra, where allele frequency of A ranges between 30-60% [17]. However, this is an evolving situation, as India goes through a ferocious second wave. It is however noted that with a median risk allele frequency of over 30%, India is an at-risk nation, with only some regions expected to have genetic protection.

The COVID-19, a viral pneumonia [21, 22], is caused by severe acute respiratory syndrome coronavirus 2 (SARS-CoV-2) [23-26]. The disease is caused by host inflammatory responses affecting the lungs and lung blood vessels [27]. Physiologically, the virus enter into the host system by attaching to angiotensin converting enzyme 2 (ACE2) receptors [28]. The viral RNA, on release, hijacks the host cell’s machinery to initiate viral replication and spread. The SARS-CoV-2 infection, triggers both innate and adaptive immune responses (both humoral and cell-mediated immunity). In the later stage when the infection spreads the epithelial-endothelial barrier integrity of lungs is compromised. It also infects the lung capillary endothelial cells, producing a rush of inflammatory response by influx of an army of immune cells like monocytes, neutrophils, macrophages infiltrating to the alveolar spaces [29]. This activates the host immune system to release the cytokines, several inflammatory cytokines (IL-1, IL-6, TNFα, and IFNγ) [30]. Interestingly, the CYP P450 enzyme activity has been involved in the inflammatory response [31, 32]. Fang *et al*. first observed a significant reduction in *CYP1A1* expression in pigs infected with *Mycoplasma hyopneumoniae* compared with the naive controls lung [33]. Later, a report validated that CYP1A1 suppress the inflammatory response caused by *M. hyopneumoniae* infection, via PPAR-signaling pathway [32]. Tian Li X *et al*. demonstrated on bacterial lipopolysaccharide (LPS) stimulation, *CYP1A1* gene was upregulated, resulting in increased production of pro-inflammatory cytokines like TNF-α and IL-6 [5]. They also suggested that this upregulation reduced the phagocytosis of the bacteria in the macrophages by decreasing the expression of SR-A, a macrophage channel crucial for phagocytosis. As a target regulated by *CYP1A1*, PPAR-γ inhibits the expression of inflammatory activating factors along with NF-κB factors [32] and increased *CYP1A1* expression is repressed by inhibition of NF-κB [34]. This indicated that *CYP1A1* expression in pulmonary macrophage is vital for host defence via regulating macrophages phagocytosis. Meanwhile, the production of pro-inflammatory cytokines drastically downregulate *CYP1A1*, disrupting the balance between the CYP enzymes, producing reactive oxygen species (ROS) thereby increasing the oxidative stress and triggering apoptosis [35, 36]. These pro-inflammatory cytokines inhibit apoptotic cell clearance in the lung, worsening the inflammation [37, 38]. Although the exact physiological role of *CYP1A1* with respect to SAR-CoV-2 has not been established, this genetic association with the current pandemic is plausible based on available data from other systems.

We note that there can be several other non-genetic factors that may increase the disease incidence. Demographic and socio-economic factors may account for COVID-19 case fatality rate (CFR) variations [39, 40]. Here, we focussed on environmental factors that may be indirectly interacting with *CYP1A1*. While there have been concerns about air pollution leading to increased COVID-19 deaths [41], the most affected nations seem to be those with the best air quality [40]. In one of our previous findings, we have shown that the A allele (rs2606345) is responsible for a 70-80% reduction in promoter activity, which would reduce enzyme activity [17]. As a caveat, induction of *CYP1A1* would be expected to be protective. This interpretation could explain counterintuitive lower mortality in regions with higher ambient air pollution, since pollutants are known to be a powerful inducer of *CYP1A1* gene expression [42]. The unexpected associations of smoking (induces *CYP1A1* gene expression [43, 44] with COVID-19 severity [45] and death [46] could also be understood in this framework. It is clearly noted that both air pollution and smoking are extremely detrimental to health and overall increase respiratory as well as non-respiratory morbidity and mortality.

To conclude, we find that *CYP1A1* alleles are associated with CAP mortality, presumably via altered xenobiotic metabolism. We speculate that gene-environment interactions governing *CYP1A1* expression may influence COVID-19 mortality. Towards this by-product of the main conclusion, we fully acknowledge there are several other factors like demographics (average age of population, population density, sex of a person, ethnic diversity, genetic variability), comorbidities (cardiovascular, cancer, and chronic respiratory diseases), socio-economic factors (GDP per capita, healthcare infrastructure), and political regime (Govt. isolation policies, social distancing, stringency index) that contribute to SARS-CoV-2 mortality and could be potential confounders. Further, the uncertainty of estimation of mortality, while the pandemic is still on, the limitations of the meta-analysis in itself, with limited population data and sample size are some major pitfalls.

## Materials and Methods

This meta-analysis was conducted as per the recommendations of the preferred reporting items for systematic reviews and meta-analyses (PRISMA) guidelines [47]. The selected studies were those in which the relationship between *CYP1A1* gene polymorphisms and risk of pneumonia disease has been evaluated. Bibliographic databases like MEDLINE (PubMed), Web of Science, and Cochrane database of systematic reviews were searched for all articles published till January 13, 2020. The keywords used to identify relevant studies were “*CYP1A1*”, “Pneumonia”, “genetic variant”, and “single nucleotide polymorphisms” using AND/ OR Boolean operators. Cross references of each study retrieved were also examined for inclusion in case they discuss the effect of *CYP1A1* genetic variant and its risk in pneumonia.

Two investigators independently reviewed each study for its inclusion in the meta-analysis. The inclusion criteria were: (1) studies conducted on human population only, (2) included the effect of *CYP1A1* genetic variant with available numeric data, (3) adopted a case-control study design (4) provided a detailed assay method. Studies (1) any other type of lung inflammation apart from pneumonia or pneumonia as a consequence of any exposure or pneumonia existing with comorbid conditions, (2) no defined diagnostic criteria for pneumonia, and (3) genotypic data not in accordance with Hardy Weinberg equilibrium were excluded.

Allele frequency data extracted for each case and control were extracted into contingency tables to calculate the odds of pneumonia in patients carrying the risk allele of the associated variants. In case of missing allele frequency data, the corresponding OR and p value were calculated from genotypic data given. The references of the retrieved articles were manually screened to identify additional studies. In case of studies where genotypic data is given allelic data is calculated to maintain a consensus across studies. The included studies and their characteristics like first author, year of publication, population, disease, genetic variant, odds ratio, genotyping method, risk allele, sample size (cases and control) and male female distribution and their quality assessment score. All the articles described some variant of pneumonia infection, one study discussing *Mycoplasma pneumoniae* infection [6], community acquired pneumonia (CAP) [8, 11, 12, 15, 48], nosocomial pneumonia (NP) [7, 49], both CAP and NP [9, 14] and relapsing pneumonia [10]. Since *M. pneumoniae* infection is the most common form of CAP, the data from this article is included of CAP cohort. Patients having frequently recurring (relapsing) pneumonia (J18, according to the ICD-10) with unspecific organism of infection, this has also been included for CAP for easy interpretation. The included cases were diagnosed by experienced professionals based on symptoms, medical histories and the clinical, radiology or laboratory results (chest X-ray, spirometry measures, etc.), and physical examination of new lung infiltrates or lower respiratory tract infection. Controls were age and gender matched healthy volunteers with no previous history of relevant infectious diseases.

Stata 16.0 (Stata Corporation, College Station, TX) [50, 51] was used to generate pooled ORs using inverse variance-weighted, fixed effects meta-analysis [52, 53]. Heterogeneity of data was evaluated using the I^2^ statistic, with I^2^ greater than 50% considered significant heterogeneity [54]. Summary ORs were represented as a point estimate and 95% confidence intervals (CIs) on a forest plot [55], and publication bias was evaluated [56-58]. The methodological assessment of all the selected articles was performed by two reviewers independently using the modified Newcastle-Ottawa Scale (NOS) for non-randomised studies [59]. The quality weight was assigned on the basis of eight categories primarily based on three broad criteria: selection of study groups; comparability of the groups; and ascertainment of either the exposure or outcome of interest for case-control, respectively. A maximum of one star was awarded for each detail present in the study for each parameter except for comparability, where a maximum of two stars can be given. A cumulative score of the number of stars obtained for each study reflected its quality. In case of conflicting scores, a consensus was reached upon discussing with another author. A study was regarded as a high-quality study when it rated six or more stars.

We performed a spatial analysis (among countries) to find the association between COVID-19 mortality with the alternate allele of rs2606345 (*CYP1A1*). We used the online resource https://ourworldindata.org/ to retrieve data on COVID-19 mortality (death per million) for each country as on May 24, 2021. We also included air pollution data obtained as a level of exposure to concentration of suspended particulate matter in air measuring less than 2.5 microns in diameter (PM_2.5_) retrieved on till May 24, 2021. The population specific allele frequency data for rs2606345 was obtained from 1000genome browser [16] on March 08, 2021. For the Indian population data, the recorded COVID-19 related death was retrieved from the ministry of health and family welfare (https://www.mohfw.gov.in/) as on 24 May 2021. The regional case fatality rate (CFR) was calculated as total number of COVID-19 related death upon a sum of total death and total patients recovered [60]. The R version 3.4.2 (R Project for Statistical Computing) was used for statistical analyses and spatial analysis plots. Initially, the data cleaning process was performed wherein we remove the missing values from the two datasets. The variable of interest includes total death per millions and SNP (rs2606345) ‘A’ allele frequency. We first estimated if COVID-19 mortality among countries varied with the allele frequency. For this, we conducted a linear regression and calculated the Pearson’s correlation coefficient. Pearson’s correlation analysis to investigate the linear or nonlinear relationship between two continuous variables. Utilizing this association, a simple linear regression model was applied to measure the relationship between mortality (dependent variable) and ‘A’ allele frequency of rs2606345 (independent variable). The goodness of fit (r^2^) was evaluated to determine the model (p< 0.05).

## Data Availability

Authors can confirm that all relevant data are included in the article and/or its supplementary information files.

## SUPPLEMENTARY MATERIALS

The Supplementary Material for this article can be found online at

## AUTHOR CONTRIBUTIONS

RK devised the concept of the review, reviewed the manuscript and supervised the overall study till final manuscript preparation. DG and SY performed the literature search independently and, reviewed articles and, extracted the data. DG generated the forest and funnel plots. PS (Priyanka Singh) performed the linear regression and calculated Pearson’s correlation and prepared all the spatial analysis images. SG helped us through the meta-analysis and running the statistical analysis through STATA. DG and PS (Pooja Singh) performed the quality assessment of the included articles and prepared the supplementary material. Upon discrepancies, RK resolved the conflict. DG and SY wrote the manuscript. ST cross-checked all the data, re-calculated the numerical values and statistical analysis. NK, SK, PRP helped in manuscript writing, reference management, and preparation of tables. RK and AA reviewed the manuscript, figures and tables. DG, SY and NK edited the manuscript. SY and NK prepared the tables. RK, YH, and BP finally revised the figures, tables and the whole manuscript. VS improved the writing of the overall manuscript. RK supervised the whole study. AA gave valuable inputs regarding the COVID-19 mortality rate and directed us to estimate the inter-ethnic difference for COVID-19 mortality. AA reviewed the manuscript. All authors have read and agreed to the published version of the manuscript.

## FUNDING

Financial support for this research work has been provided by the Council of Scientific and Industrial Research (CSIR), grant number OLP1154.

## ACKNOWLEDGEMENTS

We are highly indebted to our founder director, Prof. S.K. Brahmachari, whose vision in the field of genomics paved us a path. SY, PS (Pooja Singh), ST, NK, and PRP acknowledge CSIR, Government of India for providing their fellowships. DG, PS (Priyanka Singh) and SK acknowledges ICMR, UGC and DBT, Govt. of India, for their financial assistance, respectively. We thank the anonymous reviewers for their helpful suggestions in improving the manuscript. The authors are thankful to Dr L.E. Salnikova, V. A. Negovsky Research Institute of General Reanimatology, Russian Academy of Medical Sciences, Moscow Neurology Division, for sharing genotypic and allelic data from their patient population.

## CONFLICTS OF INTERESTS

The authors declare no conflict of interest.

## Notes

### Competing Interest Statement

The authors have declared no competing interest.

### Author Declarations

Not applicable for this study.

