## Supplementary file for "Human genetic factors associated with pneumonia susceptibility, a cue for COVID-19 mortality"

### Supplementary files:

**Supplementary files:**

1. **Table 1:** Main characteristic of studies included in meta-analysis for *CYP1A1* genetic variants association with risk of pneumonia.
2. **Table 2:** Details of quality assessment scoring for studies included in the meta-analysis based on Newcastle Ottawa Scale (NOS).
3. **Table 3:** Pooled odds ratio for allelic comparisons for studies exploring association of *CYP1A1* variants- rs2606345, rs4646903, rs1048943 in patients with risk of nosocomial pneumonia.
4. **Figure 1:** Begg’s Funnel plot for publication bias test in studies included for *CYP1A1* vaiants associated with CAP: a) rs2606345, b) rs4646903, c) rs1048943. Each point – represent a separate study for the indicated association (log) or natural logarithim of odds ratio. Horizontal line means effect size.

**Table 1:** Main characteristic of studies included in meta-analysis for *CYP1A1* genetic variants associated with risk of pneumonia

|  | **Study details** | | | | **Case** | | | | **Control** | | | |  | **Genotypic** |  | **Allelic** | | **Score** |
| --- | --- | --- | --- | --- | --- | --- | --- | --- | --- | --- | --- | --- | --- | --- | --- | --- | --- | --- |
| **No** | **Study (year) [Ref]** | **Population** | **Genotyping method** | **Disease** | **M** | **F** | **Total** | **Age (in years)** | **M** | **F** | **Total** | **Age (in years)** | **Studied *CYP1A1* variants** | **p value** | **OR (95%CI)** | **p value** | **OR (95%CI)** | **Quality** |
| 1 | Zhao J. *et* al. (2017) [6] | China | PCR Sequencing | MPP | 225 | 190 | 415 | 5.13±2.81 | 154 | 146 | 300 | 5.02± 1.63 | rs2606345 | **TT (<0.0001)** | TT 11.38(6.29-20.57) | T(0.764) | 1.07(0.68-1.66) | 7 |
| 2 | Salnikova LE *et* al.(2013) [8] | Russia | Allele specific tetra-primer PCR | CAP | 307 | 27 | 334 | 26.93±0.42 | 130 | 11 | 141 (without CAP) | 21.06±0.42 | rs2606345, rs4646903, rs1048943 | **T/T rec (3.9 × 10^-5^ )**, TT (0.093) , AA (0.188) | TT 2.40(1.59-3.64) ,TT 1.54(0.95-2.50) ,AA 0.58(0.26-1.30) | **T(<0.0001)**, **T(0.117)**, A(0.19) | T 1.90(1.41-2.55), T 1.43(0.91-2.25), A 0.59(0.27-1.3) | 5 |
|  |  |  |  |  |  |  |  |  | 286 | 28 | 314 (Healthy) | 41.65±1.03 |  | **T/T rec (1.4 × 10^-5^)**, TT(0.220), AA (0.0780) | TT 2.00(1.46-2.74) , TT 1.30(0.87-1.94), AA 0.88(0.51-1.53) | **T(0.00045)**, T(0.22), A(0.56) | T 1.5(1.20-1.9), T 1.2(0.86-1.83), A 0.68(0.60-1.3) |  |
| 3 | SalnikovaLE *et* al. (2013) [7] | Russia | PCR-CTPP | NP | 224 | 44 | 268 | 43.1±1.2 | 116 | 35 | 151 | 42.5±1.5 | rs2606345 , rs4646903, rs1048943 | TT (0.324), TT( 0.377), AA (0.88) | TT 1.23 (0.81-1.86), TT 0.79(0.47-1.32), AA 1.05 (0.50-2.22) | T(0.32), T(0.36), A(0.89) | T 1.16(0.86-1.55), T 0.79(0.48-1.29), A 1.05(0.50-2.18) | 7 |
| 4 | Salnikova LE *et* al. (2008) [9] | Russia | PCR - Genotyping | CAP | NA | NA | 99(CAP) | 30.2±13.1 | NA | NA | 160 | 21.5±5.5 | rs1048943 | **AA (0.035)** | AA 0.39(0.16-0.96) | **A (0.039)** | A 0.41(0.17-0.98) | 4 |
|  |  |  |  | NP |  |  | 57 (NP) | 48.0±14.7 |  |  |  |  |  | AA (NA) | AA 0.61(0.20-1.93) | A (NA) | A 0.63(0.20-1.92) |  |
| 5 | Korytina GF *et al.* (2005) [10] | Russia | PCR-RFLP | RP | 33 | 17 | 50 | 11.4±1.7 | 94 | 133 | 227 | 12.5±1.3 | rs1048943 | **AA (0.031)** | AA 0.25(0.08-0.72) | **A(0.009)** | A 0.25(0.099-0.67) | 6 |
| 6 | Salnikova LE *et al.* (2010) [11] | Russia | Allele specific PCR genotyping | CAP | NA | NA | 243 | NA | NA | NA | 178 | 21.53±5.49 | rs2606345 , rs4646903, rs1048943 | **TT (0.01)** | TT1.61(1.09-2.38) | **T(0.01)** | T 1.43(1.06-1.91) | 5 |
| 7 | Bold TV *et al.* (2011) [12] | Russia | Comprehensive PCR based | CAP | NA | NA | 277 (CAP) | 25.29±8.01 | NA | NA | 178 | NA | rs2606345 | **TT (0.011)** | TT1.6(1.11-2.43) | **T(0.0103)** | T 1.466(1.093-1.963) | 5 |
|  |  |  |  | NP |  |  | 158 (NP) | 43.70±17.69 |  |  |  |  |  | **TT (1 x 10^-4^)** | TT(0) | **T(<0.0001)** | T 0.48 (0.35-0.65) |  |
| 8 | SalnikovaLE. *et al.* (2013) [14] | Russia | Allele specific tetra-primer PCR | CAP | 307 | 27 | 334 | 26.9±0.8 | 130 | 11 | 141 | 29.1±0.6 | rs2606345, rs4646903, rs1048943 | **GG dom (3.9 × 10^-5^)**, CC dom (0 .086), GG dom (0.17) | TT 2.4(1.58-3.64), TT 1.53(0.94-2.50), AA 0.58(0.26-1.30) | **T(2.0 x 10^-5^)**, T(0.144), A(0.217) | T 1.90(1.41-2.55), T 1.43(0.91-2.25), A 0.59 (0.27-1.31) | 6 |
|  |  |  |  | NP | 176 | 40 | 216 | 43.0±2.0 | 83 | 22 | 105 | 41.0±1.6 | rs2606345, rs4646903, rs1048943 | GG dom (0.066), CC dom (0 .08), GG dom (0.5) | TT 1.59(0.96-2.62), TT 0.92(0.50-1.96), AA 1.34(0.57-3.11) | T(0.13), T(0.74), A (0.52) | T 1.31(0.93-1.85), T 0.91(0.52-1.59), A 1.32(0.58-3.00) |  |
| 9 | Salnikova LE.*et al.* (2013) [15] | Russia | Allele specific tetra-primer PCR | CAP | 321 | 29 | 350 | 27.2±0.8 | 343 | 89 | 432 | 30.0±0.7 | rs2606345 , rs4646903, rs1048943 | **GG dom (1.5 × 10^-6^),** CC dom(0.78), GG dom (0.52) | TT 20.3(1.52-2.71), TT 1.32 (0.92-1.89), AA 0.85(0.52-1.40) | **TT (<0.0001)**, T (0.14), A(0.54) | T 1.58(1.27-1.96), T 1.28(0.91-1.80), A 0.86(0.53-1.39) | 6 |
| 10 | Smelaya TV *et al*. (2015) [47] | Russia | Allele specific tetra-primer PCR | NP | 224 | 44 | 266^#^ | 43.1±1.2 | 116 | 35 | 150 | 42.5±1.5 | rs2606345, rs4646903, | 0.30(dom), 0.40(dom), 0.83(dom) | TT 0.81(0.53-1.22).    TT 1.26(0.75-2.11).   AA 0.94(0.44-1.99) | NA | T 0.86(0.64-1.15). T 1.25(0.77-2.04). A 0.94(0.45-1.96) | 7 |

**Bold** characters highlight important phenotypic groupings and their total counts in the respective study. M, male; F, female; PCR, polymerase chain reaction; PCR-CTPP, polymerase chain reaction- confronting two-pair primers; RFLP, restriction fragment length polymorphism; MPP, mycoplasma pneumoniae pneumonia; CAP, community acquired pneumonia; NP, nosocomial pneumonia; RP, relapsing pneumonia; OR, odds ratio; CI, confidence interval; dom, dominant model; rec, recessive model. All p values represented are uncorrected. #male/female count not given for 3 samples. Quality assessment was performed using modified NOS scale [57] the detailed scoring can be found in **Suppl. Table 2**. All the citations are as in the main manuscript file.

**Table 2:** Details of quality assessment scoring for studies included in the meta-analysis based on Newcastle Ottawa Scale (NOS)

| **Sl. No.** | **Study** | **Selection** | | | | **Comparability** | **Exposure** | | | **Cumulative score** |
| --- | --- | --- | --- | --- | --- | --- | --- | --- | --- | --- |
|  |  | Criteria 1 | Criteria 2 | Criteria 3 | Criteria 4 | Criteria 1 | Criteria 1 | Criteria 2 | Criteria 3 |  |
| 1 | Zhao J. et al., 2017 | * | * | * | * | * | * | * |  | 7 |
| 2 | Salnikova L. E. et al., 2012 | * | * | * | * |  |  | * |  | 5 |
| 3 | Salnikova L. E. et al., 2014 | * | * |  | * | ** | * | * |  | 7 |
| 4 | Salnikova L. E. et al., 2008 | * | * |  | * |  |  | * |  | 4 |
| 5 | Korytina G. F. et al., 2005 | * | * | * | * |  | * | * |  | 6 |
| 6 | Salnikova L. E. et al., 2010 | * | * |  | * |  | * | * |  | 5 |
| 7 | Bold T. V. et al., 2011 | * | * |  | * |  | * | * |  | 5 |
| 8 | Salnikova L. E. et al., 2013 | * | * |  | * | ** |  | * |  | 6 |
| 9 | Salnikova L. E. et al., 2013 | * | * |  | * | ** |  | * |  | 6 |
| 10 | Smelaya T. V. et al., 2015 | * | * | * | * | ** |  | * |  | 7 |

The Newcastle-Ottawa Scale (NOS) assesses the study quality in 3 categories: selection, comparability, and exposure. A maximum of 4 stars (*) could be given to selection items, 2 stars to comparability section and 3 stars to Exposure category.

**Table 3:** Pooled odds ratio for allelic comparisons for studies exploring association of *CYP1A1* genetic variants- rs2606345, rs4646903, rs1048943 in patients with risk of NP.

| **Gene(SNP)** | **No of study** | **Population** | **Total samples** | **NP patients** | | **Total** | **Control** | | **Total** | **OR(95%CI)** | **p value** | **I^2^** | **Model** | **Test of publication bias** |
| --- | --- | --- | --- | --- | --- | --- | --- | --- | --- | --- | --- | --- | --- | --- |
|  |  |  |  | **Risk allele present (G)** | **Risk allele absent (T)** |  | **Risk allele present (G)** | **Risk allele absent**  **(T)** |  |  |  |  |  | **Egger's test** |
| CYP1A1 (rs2606345) | 3 | Russian | 1062 | 505 | 759 | 1264 | 331 | 529 | 860 | 1.06(0.89-1.27) | 0.497 | 91.4 | R | 0.866 |
| CYP1A1 (rs1048943) | 3 | Russian | 914 | 40 | 976 | 1016 | 31 | 781 | 812 | 1.03(0.64-1.67) | 0.88 | 0 | F | NA |
| CYP1A1 (rs4646903) | 2 | Russian | 693 | 99 | 799 | 898 | 46 | 442 | 488 | 1.19(0.82-1.72) | 0.35 | 0 | F | 0.433 |

**Bold** characters highlight significantly associated alleles with respective p values. NP, nosocomial pneumonia; OR, odds ratio; CI, confidence interval; R, random effect model; F, fixed effect model.

**Figure 1:** Funnel plot for publication bias test in studies included for *CYP1A1* genetic variants associated with CAP: a) rs2606345, b) rs4646903, c) rs1048943. Each point – represent a separate study for the indicated association (log) or natural logarithim of odd ratio. Horizontal line means effect size.

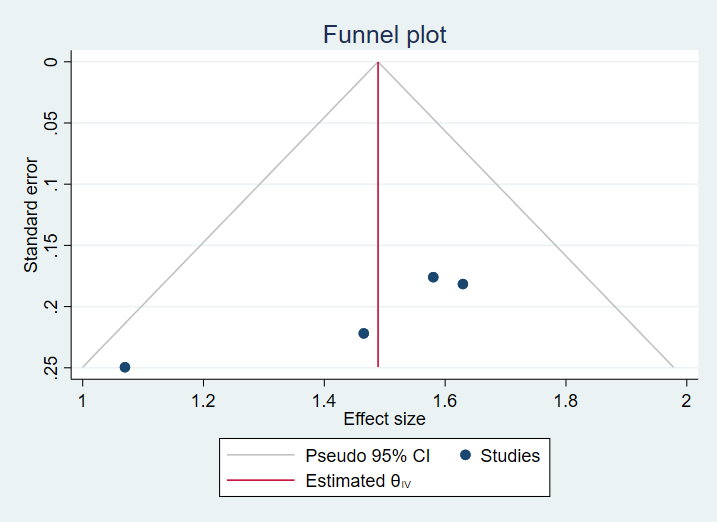
a)

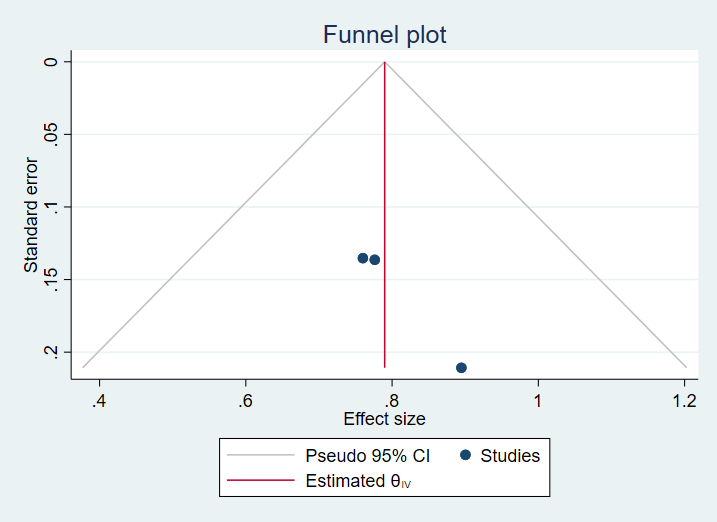

b)

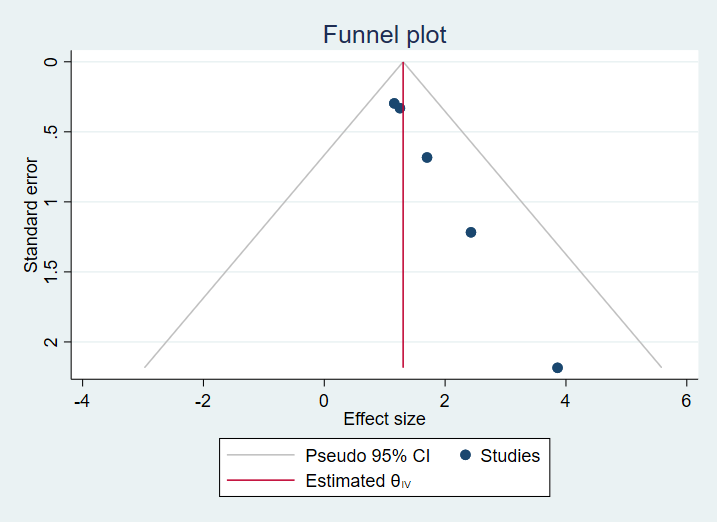

c)
